# Omicron sub-lineages BA.4/BA.5 escape BA.1 infection elicited neutralizing immunity

**DOI:** 10.1101/2022.04.29.22274477

**Authors:** Khadija Khan, Farina Karim, Yashica Ganga, Mallory Bernstein, Zesuliwe Jule, Kajal Reedoy, Sandile Cele, Gila Lustig, Daniel Amoako, Nicole Wolter, Natasha Samsunder, Aida Sivro, James Emmanuel San, Jennifer Giandhari, Houriiyah Tegally, Sureshnee Pillay, Yeshnee Naidoo, Matilda Mazibuko, Yoliswa Miya, Nokuthula Ngcobo, Nithendra Manickchund, Nombulelo Magula, Quarraisha Abdool Karim, Anne von Gottberg, Salim S. Abdool Karim, Willem Hanekom, Bernadett I. Gosnell, COMMIT-KZN Team, Richard J. Lessells, Tulio de Oliveira, Mahomed-Yunus S. Moosa, Alex Sigal

**Affiliations:** Africa Health Research Institute, Durban, South Africa; School of Laboratory Medicine and Medical Sciences, University of KwaZulu-Natal, Durban, South Africa; Centre for the AIDS Programme of Research in South Africa, Durban, South Africa; National Institute for Communicable Diseases of the National Health Laboratory Service, Johannesburg, South Africa; School of Health Sciences, College of Health Sciences, University of KwaZulu-Natal, KwaZulu-Natal, South Africa; School of Pathology, Faculty of Health Sciences, University of the Witwatersrand, Johannesburg, South Africa; Department of Medical Microbiology, University of KwaZulu-Natal, Durban, South Africa; KwaZulu-Natal Research Innovation and Sequencing Platform, Durban, South Africa; Centre for Epidemic Response and Innovation (CERI), School of Data Science and Computational Thinking, Stellenbosch University, Stellenbosch, South Africa; Department of Infectious Diseases, Nelson R. Mandela School of Clinical Medicine, University of KwaZulu-Natal, Durban, South Africa; Department of Internal Medicine, Nelson R. Mandela School of Medicine. University of Kwa-Zulu Natal; Department of Epidemiology, Mailman School of Public Health, Columbia University, New York, NY, United States; Division of Infection and Immunity, University College London, London, UK; Department of Global Health, University of Washington, Seattle, USA; Max Planck Institute for Infection Biology, Berlin, Germany

## Abstract

The SARS-CoV-2 Omicron (B.1.1.529) variant first emerged as the BA.1 sub-lineage, with extensive escape from neutralizing immunity elicited by previous infection with other variants, vaccines, or combinations of both^1,2^. Two new sub-lineages, BA.4 and BA.5, are now emerging in South Africa with changes relative to BA.1, including L452R and F486V mutations in the spike receptor binding domain. We isolated live BA.4 and BA.5 viruses and tested them against neutralizing immunity elicited to BA.1 infection in participants who were Omicron/BA.1 infected but unvaccinated (n=24) and participants vaccinated with Pfizer BNT162b2 or Johnson and Johnson Ad26.CoV.2S with breakthrough Omicron/BA.1 infection (n=15). In unvaccinated individuals, FRNT_50_, the inverse of the dilution for 50% neutralization, declined from 275 for BA.1 to 36 for BA.4 and 37 for BA.5, a 7.6 and 7.5-fold drop, respectively. In vaccinated BA.1 breakthroughs, FRNT_50_ declined from 507 for BA.1 to 158 for BA.4 (3.2-fold) and 198 for BA.5 (2.6-fold). Absolute BA.4 and BA.5 neutralization levels were about 5-fold higher in this group versus unvaccinated BA.1 infected participants. The observed escape of BA.4 and BA.5 from BA.1 elicited immunity is more moderate than of BA.1 against previous immunity^1,3^. However, the low absolute neutralization levels for BA.4 and BA.5, particularly in the unvaccinated group, are unlikely to protect well against symptomatic infection^4^.This may indicate that, based on neutralization escape, BA.4 and BA.5 have potential to result in a new infection wave.

---

We enrolled participants in November-December 2021 during the Omicron/BA.1 infection wave in South Africa (Table S1). We enrolled 24 SARS-CoV-2 infected unvaccinated participants and 15 vaccinated participants where eight were vaccinated with Pfizer BNT162b2 and seven with Johnson and Johnson Ad26.CoV.2S (Table S2). For this study we used blood samples collected a median of 23 days after symptoms onset (IQR 18-27 for vaccinated and 20-28 for unvaccinated participants), corresponding to a time post-symptoms when Omicron/BA.1 neutralizing immunity developed and plateaued^3^. As previously described (see also Table S2), infecting virus per participant was sequenced and all successfully sequenced viruses were Omicron/BA.1^3^.

We observed that FRNT_50_ in unvaccinated participants declined from 275 for BA.1 to 36 for BA.4 (7.6-fold, 95% CI 4.9-12.0, Figure 1a) and 37 for BA.5 (7.5-fold, 95% CI 4.4-12.5, Figure 1b). In contrast, in vaccinated BA.1 infected participants, FRNT_50_ declined from 507 to 158 for BA.4 (3.2-fold, 95% CI 2.3-4.4, Figure 1c) and 198 for BA.5 (2.6-fold, 95% CI 1.8-3.7, Figure 1d). Absolute BA.4 and BA.5 neutralization levels were about 5-fold higher in the vaccinated versus unvaccinated group (Figure 1d).

**Figure 1:**
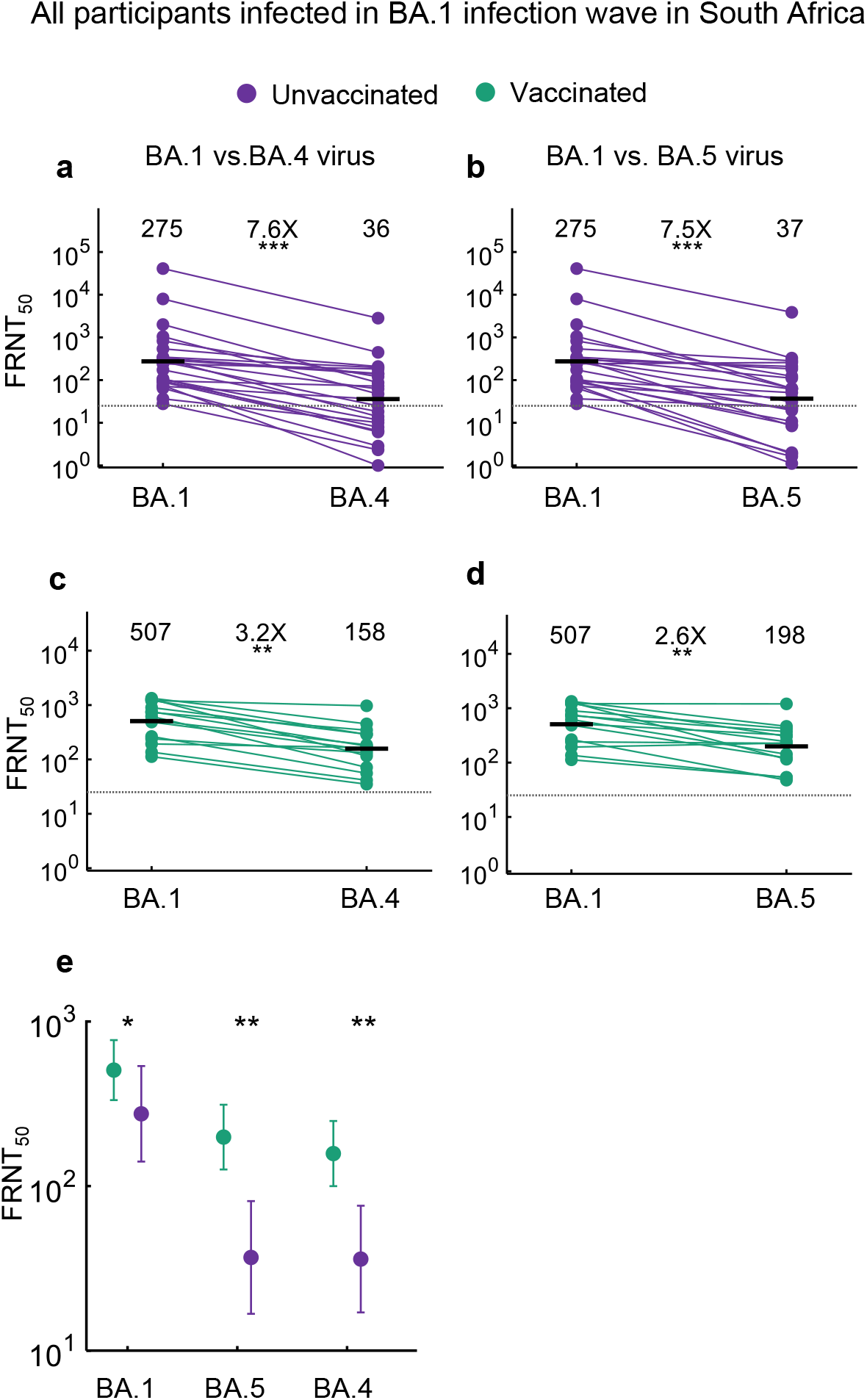
Escape of BA.4 and BA.5 from Omicron/BA.1 elicited immunity. Neutralization of BA.4 (a) or BA.5 (b) compared to BA.1 virus by immunity elicited through BA.1 infection in n=24 unvaccinated participants. Neutralization of BA.4 (c) or BA.5 (d) compared to BA.1 virus by immunity elicited through BA.1 infection in n=15 participants vaccinated either with Pfizer BNT162b2 (n=8) or Johnson and Johnson Ad26.CoV2.S (n=7). Numbers are geometric mean titer (GMT) FRNT50. Dashed line is most concentrated plasma tested. (e) Neutralization of BA.1, BA.4, and BA.5 in the n=15 Omicron/BA.1 infected vaccinated compared to the n=24 Omicron/BA.1 infected unvaccinated participants. Numbers are GMT FRNT50 and error bars are GMT 95% confidence intervals. Data is from 2 (BA.4, BA.5) or 3 (BA.1) independent experiments done on different days. p-values were determined by a two-sided Wilcoxon rank sum test and represented as *0.05-0.01, **0.01-0.001.

Neutralization capacity elicited to BA.1 infection in unvaccinated participants was low, accounting for the BA.4 and BA.5 fold-drops leading to very low residual neutralization levels. The low immunity to BA.1 is perhaps reflective of lower immunogenicity of this variant^3,5^. The vaccinated group showed about 5-fold higher neutralization capacity of BA.4 and BA.5 and should be better protected, although levels may decrease with waning. BA.4 and BA.5 show similar escape. This is not surprising given that sequence differences between BA.4 and BA.5 are outside spike.

The escape we observe is substantially higher than what we detected for BA.2, where it was slight and non-significant^3^. Given the higher escape and especially the low residual neutralization in the unvaccinated group, we speculate that a BA.4/BA.5 infection wave is a strong possibility. However, vaccination increase neutralization capacity against these emerging variants and would likely offer good protection against severe disease.

## Materials and methods

### Informed consent and ethical statement

Blood samples were obtained after written informed consent from adults with PCR-confirmed SARS-CoV-2 infection who were enrolled in a prospective cohort study at the Africa Health Research Institute approved by the Biomedical Research Ethics Committee at the University of KwaZulu–Natal (reference BREC/00001275/2020). The Omicron/BA.1 and BA.4 was isolated from a residual swab sample with SARS-CoV-2 isolation from the sample approved by the University of the Witwatersrand Human Research Ethics Committee (HREC) (ref. M210752). The sample to isolate Omicron/BA.5 was collected after written informed consent as part of the COVID-19 transmission and natural history in KwaZulu-Natal, South Africa: Epidemiological Investigation to Guide Prevention and Clinical Care in the Centre for the AIDS Programme of Research in South Africa (CAPRISA) study and approved by the Biomedical Research Ethics Committee at the University of KwaZulu–Natal (reference BREC/00001195/2020, BREC/00003106/2021).

### Data availability statement

Sequences of outgrown Omicron sub-lineages have been deposited in GISAID with accession EPI_ISL_7886688 (Omicron/BA.1), EPI_ISL_12268495.2 (Omicron/BA.4), EPI_ISL_12268493.2 (Omicron/BA.5). Raw images of the data are available upon reasonable request.

### Reagent availability statement

Virus isolates and cell line are available from the corresponding author. A Biosafety Level 3 facility is required for laboratories receiving live SARS-CoV-2.

## Author contributions

AS and KK conceived the study and designed the study and experiments. AvG, QAK, SSAK, GL, ASi, and NS identified and provided virus samples. M-YSM, FK, BIG, MB, KK, TN, MM, NM, ZM, NN, YM, ZJ, KR, and YG set up and managed the cohort and cohort data. KK, ZJ, KR, SC, HT, JES, YG, JG, YR, AK, DA, and JB performed experiments and sequence analysis with input from AS, TdO, RJL, and JNB. AS, KK, FK, RM, and YR interpreted data with input from M-YSM, GG, SSAK, WH, TdO, RJL. AS and KK prepared the manuscript with input from all authors.

## Whole-genome sequencing, genome assembly and phylogenetic analysis

RNA was extracted on an automated Chemagic 360 instrument, using the CMG-1049 kit (Perkin Elmer, Hamburg, Germany). The RNA was stored at -80°C prior to use. Libraries for whole genome sequencing were prepared using either the Oxford Nanopore Midnight protocol with Rapid Barcoding or the Illumina COVIDseq Assay. For the Illumina COVIDseq assay, the libraries were prepared according to the manufacturer’s protocol. Briefly, amplicons were tagmented, followed by indexing using the Nextera UD Indexes Set A. Sequencing libraries were pooled, normalized to 4 nM and denatured with 0.2 N sodium acetate. An 8 pM sample library was spiked with 1% PhiX (PhiX Control v3 adaptor-ligated library used as a control). We sequenced libraries on a 500-cycle v2 MiSeq Reagent Kit on the Illumina MiSeq instrument (Illumina). On the Illumina NextSeq 550 instrument, sequencing was performed using the Illumina COVIDSeq protocol (Illumina Inc, USA), an amplicon-based next-generation sequencing approach. The first strand synthesis was carried using random hexamers primers from Illumina and the synthesized cDNA underwent two separate multiplex PCR reactions. The pooled PCR amplified products were processed for tagmentation and adapter ligation using IDT for Illumina Nextera UD Indexes. Further enrichment and cleanup was performed as per protocols provided by the manufacturer (Illumina Inc). Pooled samples were quantified using Qubit 3.0 or 4.0 fluorometer (Invitrogen Inc.) using the Qubit dsDNA High Sensitivity assay according to manufacturer’s instructions. The fragment sizes were analyzed using TapeStation 4200 (Invitrogen). The pooled libraries were further normalized to 4nM concentration and 25 μL of each normalized pool containing unique index adapter sets were combined in a new tube. The final library pool was denatured and neutralized with 0.2N sodium hydroxide and 200 mM Tris-HCL (pH7), respectively. 1.5 pM sample library was spiked with 2% PhiX. Libraries were loaded onto a 300-cycle NextSeq 500/550 HighOutput Kit v2 and run on the Illumina NextSeq 550 instrument (Illumina, San Diego, CA, USA). For Oxford Nanopore sequencing, the Midnight primer kit was used as described by Freed and Silander55. cDNA synthesis was performed on the extracted RNA using LunaScript RT mastermix (New England BioLabs) followed by gene-specific multiplex PCR using the Midnight Primer pools which produce 1200bp amplicons which overlap to cover the 30-kb SARS-CoV-2 genome. Amplicons from each pool were pooled and used neat for barcoding with the Oxford Nanopore Rapid Barcoding kit as per the manufacturer’s protocol. Barcoded samples were pooled and bead-purified. After the bead clean-up, the library was loaded on a prepared R9.4.1 flow-cell. A GridION X5 or MinION sequencing run was initiated using MinKNOW software with the base-call setting switched off. We assembled paired-end and nanopore.fastq reads using Genome Detective 1.132 (https://www.genomedetective.com) which was updated for the accurate assembly and variant calling of tiled primer amplicon Illumina or Oxford Nanopore reads, and the Coronavirus Typing Tool56. For Illumina assembly, GATK HaploTypeCaller -- min-pruning 0 argument was added to increase mutation calling sensitivity near sequencing gaps. For Nanopore, low coverage regions with poor alignment quality (<85% variant homogeneity) near sequencing/amplicon ends were masked to be robust against primer drop-out experienced in the Spike gene, and the sensitivity for detecting short inserts using a region-local global alignment of reads, was increased. In addition, we also used the wf_artic (ARTIC SARS-CoV-2) pipeline as built using the nextflow workflow framework57. In some instances, mutations were confirmed visually with .bam files using Geneious software V2020.1.2 (Biomatters). The reference genome used throughout the assembly process was NC_045512.2 (numbering equivalent to MN908947.3). For lineage classification, we used the widespread dynamic lineage classification method from the ‘Phylogenetic Assignment of Named Global Outbreak Lineages’ (PANGOLIN) software suite (https://github.com/hCoV-2019/pangolin).

## Cells

Vero E6 cells (originally ATCC CRL-1586, obtained from Cellonex in South Africa) were propagated in complete growth medium consisting of Dulbecco’s Modified Eagle Medium (DMEM) with 10% fetal bovine serum (Hyclone) containing 10mM of HEPES, 1mM sodium pyruvate, 2mM L-glutamine and 0.1mM nonessential amino acids (Sigma-Aldrich). Vero E6 cells were passaged every 3–4 days. The H1299-E3 cell line (H1299 originally from ATCC as CRL-5803) was propagated in growth medium consisting of complete Roswell Park Memorial Institute (RPMI) 1640 medium with 10% fetal bovine serum containing 10mM of HEPES, 1mM sodium pyruvate, 2mM L-glutamine and 0.1mM nonessential amino acids. Cells were passaged every second day. The H1299-E3 (H1299-ACE2, clone E3) cell line was derived from H1299 as described in our previous work^1,6^.

## Virus expansion

All work with live virus was performed in Biosafety Level 3 containment using protocols for SARS-CoV-2 approved by the Africa Health Research Institute Biosafety Committee. ACE2-expressing H1299-E3 cells were seeded at 4.5 × 10^5^ cells in a 6 well plate well and incubated for 18–20 h. After one DPBS wash, the sub-confluent cell monolayer was inoculated with 500 μL universal transport medium diluted 1:1 with growth medium filtered through a 0.45-μm filter. Cells were incubated for 1 h. Wells were then filled with 3 mL complete growth medium. After 4 days of infection (completion of passage 1 (P1)), cells were trypsinized, centrifuged at 300 rcf for 3 min and resuspended in 4 mL growth medium. Then all infected cells were added to Vero E6 cells that had been seeded at 2 × 10^5^ cells per mL, 20mL total, 18–20 h earlier in a T75 flask for cell-to-cell infection. The coculture of ACE2-expressing H1299-E3 and Vero E6 cells was incubated for 1 h and the flask was filled with 20 mL of complete growth medium and incubated for 4 days. The viral supernatant from this culture (passage 2 (P2) stock) was used for experiments.

## Live virus neutralization assay

H1299-E3 cells were plated in a 96-well plate (Corning) at 30,000 cells per well 1 day pre-infection. Plasma was separated from EDTA-anticoagulated blood by centrifugation at 500 rcf for 10 min and stored at -80 °C. Aliquots of plasma samples were heat-inactivated at 56 °C for 30 min and clarified by centrifugation at 10,000 rcf for 5 min. Virus stocks were used at approximately 50-100 focus-forming units per microwell and added to diluted plasma. Antibody–virus mixtures were incubated for 1 h at 37 °C, 5% CO_2_. Cells were infected with 100 μL of the virus–antibody mixtures for 1 h, then 100 μL of a 1X RPMI 1640 (Sigma-Aldrich, R6504), 1.5% carboxymethylcellulose (Sigma-Aldrich, C4888) overlay was added without removing the inoculum. Cells were fixed 18 h post-infection using 4% PFA (Sigma-Aldrich) for 20 min. Foci were stained with a rabbit anti-spike monoclonal antibody (BS-R2B12, GenScript A02058) at 0.5 μg/mL in a permeabilization buffer containing 0.1% saponin (Sigma-Aldrich), 0.1% BSA (Sigma-Aldrich) and 0.05% Tween-20 (Sigma-Aldrich) in PBS. Plates were incubated with primary antibody overnight at 4 °C, then washed with wash buffer containing 0.05% Tween-20 in PBS. Secondary goat anti-rabbit HRP conjugated antibody (Abcam ab205718) was added at 1 μg/mL and incubated for 2 h at room temperature with shaking. TrueBlue peroxidase substrate (SeraCare 5510-0030) was then added at 50 μL per well and incubated for 20 min at room temperature. Plates were imaged in an ImmunoSpot Ultra-V S6-02-6140 Analyzer ELISPOT instrument with BioSpot Professional built-in image analysis (C.T.L).

## Statistics and fitting

All statistics and fitting were performed using custom code in MATLAB v.2019b. Neutralization data were fit to:

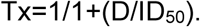

Here Tx is the number of foci normalized to the number of foci in the absence of plasma on the same plate at dilution D and ID_50_ is the plasma dilution giving 50% neutralization. FRNT_50_= 1/ID_50_. Values of FRNT_50_ <1 are set to 1 (undiluted), the lowest measurable value. We note that the most concentrated plasma dilution was 1:25 and therefore FRNT_50_ < 25 were extrapolated. To calculate confidence intervals, FRNT_50_ or fold-change in FRNT_50_ per participant was log transformed and arithmetic mean plus 2 std and arithmetic mean minus 2 std were calculated for the log transformed values. These were exponentiated to obtain the upper and lower 95% confidence intervals on the geometric mean FRNT_50_ or the fold-change in FRNT_50_ geometric means.

## Acknowledgements

This study was supported by the Bill and Melinda Gates award INV-018944 (AS), National Institutes of Health award R01 AI138546 (AS), and South African Medical Research Council Award 6084COAP2020 (AS). The funders had no role in study design, data collection and analysis, decision to publish, or preparation of the manuscript.

## COMMIT-KZN Team

Rohen Harrichandparsad^24^, Kobus Herbst^1,25^, Prakash Jeena^26^, Thandeka Khoza^1^, Henrik Kløverpris^1,27^, Alasdair Leslie^1,11^, Rajhmun Madansein^28^, Nombulelo Magula^29^, Nithendra Manickchund^13^, Mohlopheni Marakalala^1,11^, Matilda Mazibuko^1^, Mosa Moshabela^30^, Ntombifuthi Mthabela^1^, Kogie Naidoo^8^, Zaza Ndhlovu^1, 31^, Thumbi Ndung’u^1,16,31,32^, Nokuthula Ngcobo^1^, Kennedy Nyamande^33^, Vinod Patel^34^, Theresa Smit^1^, Adrie Steyn^1,35^ & Emily Wong^1,35^.

^24^Department of Neurosurgery, University of KwaZulu-Natal, Durban, South Africa. ^25^South African Population Research Infrastructure Network, Durban, South Africa. ^26^Department of Paediatrics and Child Health, University of KwaZulu-Natal, Durban, South Africa. ^27^Department of Immunology and Microbiology, University of Copenhagen, Copenhagen, Denmark. ^28^Department of Cardiothoracic Surgery, University of KwaZulu-Natal, Durban, South Africa. ^29^Department of Medicine, King Edward VIII Hospital and University of KwaZulu Natal, Durban, South Africa.^30^College of Health Sciences, University of KwaZulu-Natal, Durban, South Africa. ^31^Ragon Institute of MGH, MIT and Harvard, Boston, USA. ^32^HIV Pathogenesis Programme, The Doris Duke Medical Research Institute, University of KwaZulu-Natal, Durban, South Africa. ^33^Department of Pulmonology and Critical Care, University of KwaZulu-Natal, Durban, South Africa. ^34^Department of Neurology, University of KwaZulu-Natal, Durban, South Africa.^35^Division of Infectious Diseases, University of Alabama at Birmingham, USA.

**Table S1:** Summary of participant details.

|  | All<br>39 | Vaccinated<br>15 (38%) | Unvaccinated<br>24 (62%) |
| --- | --- | --- | --- |
| Age | 35 (27-55) | 37 (32-60) | 31.5 (26-49) |
| Female | 25 (64%) | 9 (60%) | 16 (67%) |
| Vaccination to symptom onset (days) | - | 135 (111-195) | - |
| Symptom onset to collection (days) | 23 (19-27) | 23 (18-27) | 23 (20-28) |
| Required supp. O<sub>2</sub> | 7 (18%) | 2 (13%) | 5 (21%) |
| Hospitalized | 27 (69%) | 8 (53%) | 19 (79%) |
| Duration of hospitalization (days) | 7 (3-11) | 3.5 (2.5-14.5) | 8 (3-11) |

**Table S2:**
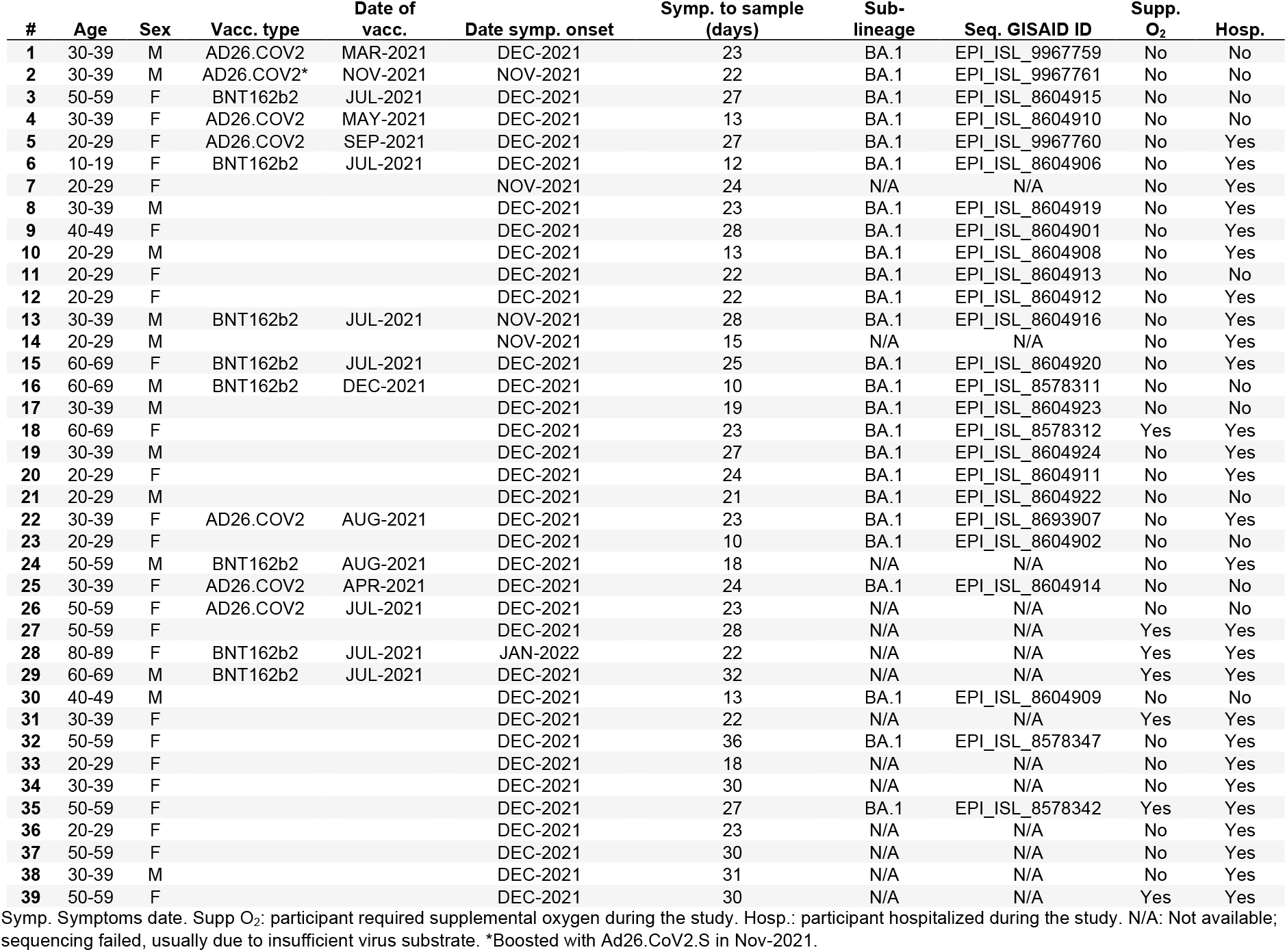
Participant characteristics and vaccination status.

| # | Age | Sex | Vacc. type | Date of<br>vacc. | Date symp. onset | Symp. to sample<br>(days) | Sub-lineage | Seq. GISAID ID | Supp.<br>O<sub>2</sub> | Hosp. |
| --- | --- | --- | --- | --- | --- | --- | --- | --- | --- | --- |
| 1 | 30-39 | M | AD26.COV2 | MAR-2021 | DEC-2021 | 23 | BA.1 | EPI_ISL_9967759 | No | No |
| 2 | 30-39 | M | AD26.COV2* | NOV-2021 | NOV-2021 | 22 | BA.1 | EPI_ISL_9967761 | No | No |
| 3 | 50-59 | F | BNT162b2 | JUL-2021 | DEC-2021 | 27 | BA.1 | EPI_ISL_8604915 | No | No |
| 4 | 30-39 | F | AD26.COV2 | MAY-2021 | DEC-2021 | 13 | BA.1 | EPI_ISL_8604910 | No | No |
| 5 | 20-29 | F | AD26.COV2 | SEP-2021 | DEC-2021 | 27 | BA.1 | EPI_ISL_9967760 | No | Yes |
| 6 | 10-19 | F | BNT162b2 | JUL-2021 | DEC-2021 | 12 | BA.1 | EPI_ISL_8604906 | No | Yes |
| 7 | 20-29 | F |  |  | NOV-2021 | 24 | N/A | N/A | No | Yes |
| 8 | 30-39 | M |  |  | DEC-2021 | 23 | BA.1 | EPI_ISL_8604919 | No | Yes |
| 9 | 40-49 | F |  |  | DEC-2021 | 28 | BA.1 | EPI_ISL_8604901 | No | Yes |
| 10 | 20-29 | M |  |  | DEC-2021 | 13 | BA.1 | EPI_ISL_8604908 | No | Yes |
| 11 | 20-29 | F |  |  | DEC-2021 | 22 | BA.1 | EPI_ISL_8604913 | No | No |
| 12 | 20-29 | F |  |  | DEC-2021 | 22 | BA.1 | EPI_ISL_8604912 | No | Yes |
| 13 | 30-39 | M | BNT162b2 | JUL-2021 | NOV-2021 | 28 | BA.1 | EPI_ISL_8604916 | No | Yes |
| 14 | 20-29 | M |  |  | NOV-2021 | 15 | N/A | N/A | No | Yes |
| 15 | 60-69 | F | BNT162b2 | JUL-2021 | DEC-2021 | 25 | BA.1 | EPI_ISL_8604920 | No | Yes |
| 16 | 60-69 | M | BNT162b2 | DEC-2021 | DEC-2021 | 10 | BA.1 | EPI_ISL_8578311 | No | No |
| 17 | 30-39 | M |  |  | DEC-2021 | 19 | BA.1 | EPI_ISL_8604923 | No | No |
| 18 | 60-69 | F |  |  | DEC-2021 | 23 | BA.1 | EPI_ISL_8578312 | Yes | Yes |
| 19 | 30-39 | M |  |  | DEC-2021 | 27 | BA.1 | EPI_ISL_8604924 | No | Yes |
| 20 | 20-29 | F |  |  | DEC-2021 | 24 | BA.1 | EPI_ISL_8604911 | No | Yes |
| 21 | 20-29 | M |  |  | DEC-2021 | 21 | BA.1 | EPI_ISL_8604922 | No | No |
| 22 | 30-39 | F | AD26.COV2 | AUG-2021 | DEC-2021 | 23 | BA.1 | EPI_ISL_8693907 | No | Yes |
| 23 | 20-29 | F |  |  | DEC-2021 | 10 | BA.1 | EPI_ISL_8604902 | No | No |
| 24 | 50-59 | M | BNT162b2 | AUG-2021 | DEC-2021 | 18 | N/A | N/A | No | Yes |
| 25 | 30-39 | F | AD26.COV2 | APR-2021 | DEC-2021 | 24 | BA.1 | EPI_ISL_8604914 | No | No |
| 26 | 50-59 | F | AD26.COV2 | JUL-2021 | DEC-2021 | 23 | N/A | N/A | No | No |
| 27 | 50-59 | F |  |  | DEC-2021 | 28 | N/A | N/A | Yes | Yes |
| 28 | 80-89 | F | BNT162b2 | JUL-2021 | JAN-2022 | 22 | N/A | N/A | Yes | Yes |
| 29 | 60-69 | M | BNT162b2 | JUL-2021 | DEC-2021 | 32 | N/A | N/A | Yes | Yes |
| 30 | 40-49 | M |  |  | DEC-2021 | 13 | BA.1 | EPI_ISL_8604909 | No | No |
| 31 | 30-39 | F |  |  | DEC-2021 | 22 | N/A | N/A | Yes | Yes |
| 32 | 50-59 | F |  |  | DEC-2021 | 36 | BA.1 | EPI_ISL_8578347 | No | Yes |
| 33 | 20-29 | F |  |  | DEC-2021 | 18 | N/A | N/A | No | Yes |
| 34 | 30-39 | F |  |  | DEC-2021 | 30 | N/A | N/A | No | Yes |
| 35 | 50-59 | F |  |  | DEC-2021 | 27 | BA.1 | EPI_ISL_8578342 | Yes | Yes |
| 36 | 20-29 | F |  |  | DEC-2021 | 23 | N/A | N/A | No | Yes |
| 37 | 50-59 | F |  |  | DEC-2021 | 30 | N/A | N/A | No | Yes |
| 38 | 30-39 | M |  |  | DEC-2021 | 31 | N/A | N/A | No | Yes |
| 39 | 50-59 | F |  |  | DEC-2021 | 30 | N/A | N/A | Yes | Yes |

Symp.: Symptoms date. Supp O<sub>2</sub>: participant required supplemental oxygen during the study. Hosp.: participant hospitalized during the study. N/A: Not available; sequencing failed, usually due to insufficient virus substrate. \*Boosted with Ad26.CoV2.S in Nov-2021.

